# HIV self-testing and its associated factors among young university students: A cross-sectional study in a Public University in Uganda

**DOI:** 10.1101/2024.06.18.24309140

**Authors:** Winnie Nambi, Rose Chalo Nabirye, Gloria Nakato, Mary Aleni, Joshua Epuitai

## Abstract

**Introduction:** Uptake of HIV testing services is sub-optimal among young adults. HIV self-testing offers opportunities to improve uptake of HIV testing services among young adults. The study was conducted to determine preferences of HIV testing, uptake of HIV self-testing and the factors associated with HIV-self testing among young adults.

**Methods:** The study was a descriptive cross-sectional design. A self-administered questionnaire for data collection. We interviewed 384 university students to collect information about HIV testing and their preferences. Logistic regression analysis was used to determine the factors associated with HIV self-testing. The study obtained ethical clearance from the Mbale Regional Referral Hospital Research and Ethics Committee.

**Results:** The median age of the participants was 24 (interquartile range: 22-28). Uptake of HIV self-testing was 55% among young adults. Similarly, 56% of the young adults preferred HIV self-testing over facility-based HIV testing in the future. Privacy (46%), confidentiality (7%) and convenience (32%) were the most common reasons given for preferring HIV self-testing. Participants who preferred to test in the healthcare facility cited preference for counselling services (21%) and the perception that the HIV test results from the healthcare facility were more accurate (37%), credible and trustworthy (21%). HIV self-testing was significantly associated with several factors: increasing age (95% CI: 0.89 (0.80-0.98)), fourth year of study (95%CI: 2.90 (1.01-8.31), students of anaesthesia program (95% CI: 0.40 (0.17-0.95), Muslim religion (95% CI: 0. 07 (0.01-0.41), consistent use of condoms (95% CI: 0.36 (0.15-0.88), and having multiple sexual partners (95% CI: 3.22 (1.49-7.00).

**Conclusion:** Young adults preferred HIV self-testing over provider-based testing in the health facility. This preference was related to privacy, confidentiality and convenience. Addressing concerns about accuracy of test results in HIV self-testing and need for counselling services may improve uptake of HIV self-testing among young adults.

## Background

In Uganda, an estimated 1.4 million people are living with Human Immunodeficiency Virus (HIV) [1]. Young adults, especially adolescent girls, are disproportionately affected with a high risk of acquiring new HIV infections [2]. In 2023, an estimated 52,000 new HIV infections were reported in Uganda of which 36% were among people between the age of 15-24 years [1]. The AIDS- related deaths in Uganda were estimated at about 17,000 annually [1]. The 95-95-95 strategy is critical in eliminating HIV transmission by 2030 as in the sustainable development goals [3]. This strategy involves ensuring that 95% of people living with HIV (PLHIV) are aware of their HIV status, 95% of the PLHIV are on highly active antiretroviral therapy (HAART), and 95% of PLHIV on HAART have achieved viral load suppression [4].

HIV testing enables individuals to adopt prevention practices that reduce HIV transmission [4]. This ensures that PLHIV are aware of their status, thus facilitating early initiation on HAART [4]. Consequently, HIV testing helps improve quality of life for PLHIV and it also reduces AIDS related mortality [5]. Despite the critical role of HIV testing in reducing the burden of HIV, it is still suboptimal, especially among young adults [6]. In Nigeria, only 24% of adolescents and young adults had ever tested for HIV [5]. Furthermore, HIV testing in sub-Saharan Africa was least likely among young adults who were living with HIV, and therefore, were unaware of their HIV positive status [6]. HIV testing rates in Uganda were lowest among young adults 15-24 years than any other age groups [7].

The traditional HIV testing services, especially voluntary counselling and testing and provider-initiated HIV counselling and testing, have not significantly improved uptake of HIV testing services in sub-Saharan Africa [6]. Stigma, provider bias, lack of access, and concerns of privacy have affected uptake of HIV services among young adults [5,6]. The new HIV testing technologies and approaches such as HIV self-testing offers opportunities to improve uptake of HIV testing services [6]. HIV self-testing (HIVST) can occur in different forms where individuals can test at home, using oral or blood testing kits, and can access counselling services from trained healthcare professionals over the phone [8]. Individuals with positive HIV tests results after HIV home self-testing can be linked to counselling and receive further care in the healthcare facilities [9].

Young adults tend to have higher preference and acceptability of HIVST options [8,10]. In Nigeria, the majority (49%) of young adults preferred blood-based HIVST than the 44% who preferred facility-based HIV testing and oral HIVST [8]. In Uganda, 95% of university female students were willing to use HIVST [11]. HIVST addresses youth-specific barriers inherent in the traditional HIV testing services as it fosters respect, privacy, convenience, confidentiality and comfort [10,12]. In Uganda, a limited number of studies, to the best of our knowledge, have explored HIVST preferences among young adults [11,13]. Therefore, this study was conducted to determine the preferences for HIVST, uptake of HIVST and its associated factors associated among young adults in a public University.

## Methods and materials

### Study design and setting

We used a cross-sectional study to determine the HIVST preferences and practices among young adults. The study was conducted in Busitema University, Faculty of Health Sciences. This public university attracts a large number of students from different socioeconomic, cultural, and religious backgrounds. The Faculty of Health Sciences is located in Eastern Uganda, where it offers undergraduate and postgraduate programs. The undergraduate programs are in nursing, anaesthesia, medicine and surgery, while postgraduate programs are in the area of public health and master of medicine programs.

### Study population

The study population was comprised of young university students in the Faculty of Health Science who were pursuing clinical disciplines. We used consecutive sampling to select the participants for the study. The sample size, determined using the Kish-Leslie formular, was 384. This represented 85% of the student population in the Faculty of Health Sciences.

### Data collection tool

We used a self-administered questionnaire for data collection. Data was collected between 01^st^ November 2022 to 31^st^ December 2022. Participants filled the questionnaires designed on the Kobo toolbox. The questionnaire had sections for socio-demographic, sexual behaviours, and HIV testing practices (Appendix 1). The questionnaire had an open-ended question on the challenges that participants face while seeking HIV testing services, and the reasons for preferring HIVST or provider-based testing in the facilities.

### Study variables

The dependent variable in the study was HIV self-testing which was considered in individuals who tested themselves at home in the first sexual encounter with a new or current partner. Additionally, participants were asked about their preferences for HIV testing (self-testing at home or healthcare provider testing in the healthcare facility) if they were to enter into a new relationship in the future.

The independent variables included age, gender, marital status, employment status (employed/unemployed), year of study, program of study, religion, sexual risk behaviors, and HIV testing practices. The sexual risk behaviors included number of sexual partners in the last 12 months, age at sex debut, inconsistent use of condoms, non-disclosure of HIV status, broken condoms, and whether they were sexually active. HIV testing practices investigated were ever testing for HIV, number of times HIV was tested, and HIV testing at the last sexual encounter.

### Data analysis

Data was analysed using STATA version 15 software. The open-ended questions on reasons for preferring self-testing or provider-based testing were coded and labelled into categories. Categorical variables were analysed through use of frequencies and percentages, while median and interquartile range was used to describe continuous variables not normally distributed. Logistic regression at bivariate and multivariate level was used to determine the factors associated with HIV self-testing among university students. The probability value (p-value) of less than 5% indicated significant association, while the 95% confidence level (CI) was used to infer the magnitude of association. P-values which were less than 0.2 at the bivariate level and variables relevant to HIV self-testing were included in the multivariate logistic regression model. Factors associated with HIV self-testing were those that were significant at the p-value of less than 0.05 at the multivariate logistic regression.

### Ethical consideration

The Mbale Regional Referral Hospital (MRRH) Research and Ethics Committee (MRRH-2022-212) provided ethical clearance for the study. A written informed consent was obtained from all the study participants. Privacy was observed through sending participants an anonymised link to the questionnaire instead of the face-to-face interview where information from the participants could be identified. The filled information was confidential, while measures were put in place to maintain confidentiality.

## Results

### Description of study participants

The median age of the participants was 24 (interquartile range of 22-28 years), while the mean age at sex debut was 19.6 (standard deviation of 2.56) (Table 1). More than a half of the participants had two or more partners in the last year (55%), while 61% of the participants had sex with a partner of unknown HIV status.

**Table 1:** Description of the study participants.

| Variable | Frequency (percent)<br>N=384 |
| --- | --- |
| **Age (n=384)** |  |
| Median (interquartile range) | 24 (22-28) |
| **Gender (n=384)** |  |
| Male | 230 (59.9) |
| Female | 154 (40.1) |
| **Marital status (n=384)** |  |
| Single | 242 (63.0) |
| Married/cohabiting | 142 (37.0) |
| **Year of study** |  |
| Year 1 | 64 (16.7) |
| Year 2 | 92 (24.0) |
| Year 3 | 62 (16.1) |
| Year 4 | 90 (23.4) |
| Year 5 | 76 (19.8) |
| **Program of study** |  |
| Bachelor of Nursing | 71 (18.5) |
| Bachelor of Anaesthesia | 70 (18.2) |
| Bachelor of Medicine and Surgery | 243 (63.3) |
| **Employment** |  |
| Unemployed | 282 (73.4) |
| Employed | 102 (26.6) |
| **Religion** |  |
| Christian | 362 (94.3) |
| Moslem | 22 (5.7) |
| **Are you in any sexual relationship?** |  |
| No | 65 (16.9) |
| Yes | 319 (83.1) |
| **Age at sex debut** |  |
| Mean (standard deviation) | 19.6 (2.56) |
| **Knows the HIV status of the partner in the first sexual encounter (n=320)** |  |
| No | 168 (52.5) |
| Yes | 152 (47.5) |
| **Number of partners in the past year (n=320)** |  |
| 1 | 146 (45.6) |
| 2 or more | 174 (54.4) |
| **HIV high risky behaviours** |  |
| Inconsistent use of condoms | 274 (71.4) |
| Broken condoms | 175 (45.6) |
| Multiple sexual partners | 171 (44.5) |
| Sex with a person with unknown status | 234 (60.9) |
| Not disclosing the HIV status | 109 (28.4) |

### HIV testing practices among young adults

Almost all the study participants had ever tested for HIV (table 2). The majority (80%, n=258) of the participants tested for HIV in the first sexual encounter with a new partner. Among participants tested for HIV, 55% (144/260) of them opted for HIV self-testing. The challenges associated with HIV testing were it being expensive or cost of testing, unavailability and inaccessibility of the test kits or services.

**Table 2:** HIV testing practices among young adults.

| **Variable** | **Frequency,**<br>**n=320**<br>**(percentage)** |
| --- | --- |
| **Ever tested for HIV** |  |
| Yes | 375 (97.7) |
| No | 9 (2.3) |
| **Number of times of HIV testing (n=349)** |  |
| Once | 105 (30.1) |
| Multiple times | 244 (69.9) |
| **HIV testing in the last sexual encounter** |  |
| Yes | 206 (64.4) |
| No | 73 (22.8) |
| Not applicable (status known for both of us) | 41 (12.8) |
| **Tested for HIV in the first sexual encounter with a new partner (n=324)** |  |
| Yes | 258 (79.6) |
| No | 66 (20.4) |
| **Place of HIV testing in the first sexual encounter with a new partner (N=260)** |  |
| Home/self-testing | 144 (55.4) |
| Health facility/healthcare provider testing | 116 (44.6) |
| **Challenges of HIV testing** |  |
| HIV testing kits unavailable | 226 (58.8) |
| HIV testing expensive | 120 (31.2) |
| Unable to access drug store/pharmacy | 10 (2.6) |
| Unable to test myself | 28 (7.3) |

### Preferences for HIV testing options

Similar to the higher proportion of young adults who self-tested for HIV at home, more than a half of the participants (56%, n=213) preferred to test by themselves at home if they were to enter into new sexual relationships in the future (table 3). Privacy and convenience were the most common reasons given for preferring HIV-self testing. Individuals who preferred provider-based HIV testing in the health facilities did so because of concerns of accuracy of test results and the need for pre-test and post-test counselling.

**Table 3:** Preferences of HIV testing options.

| Variable | Frequency, n=320<br>(percentage) |
| --- | --- |
| **Would you wish to test for HIV in a new relationship? (N=384)** |  |
| Yes | 376 (97.9) |
| Undecided | 8 (2.1) |
| **Preferred type of HIV testing (=376)** |  |
| Home/self-testing | 213 (56.6) |
| Health facility/healthcare provider | 163 (43.4) |
| **Reasons for preferring HIV self-testing (n=238)\*\*\*** |  |
| Privacy | 110 (46.2) |
| Convenience | 76 (31.9) |
| Confidentiality | 16 (6.7) |
| Other reasons* | 36 (15.1) |
| **Reasons for preferring health facility (n=174)\*\*\*** |  |
| Proper testing and accurate results | 65 (37.3) |
| Trustworthy/credible/reliable results | 37 (21.3) |
| Pre and post-test counselling | 36 (20.7) |
| Needs to be tested by healthcare provider | 16 (9.2) |
| Safety of HIV testing | 12 (6.9) |
| Other reasons** | 8 (4.6) |
| **Preference for pre- and post-testing counselling (n=384)** |  |
| Yes | 331 (86.2) |
| No | 53 (13.8) |

\*Other reasons (able to test on their own, saves time, cheap and mistrust of healthcare workers)
\*\*Other reasons (confidentiality, convenience, easier and cannot do self-test)
\*\*\*multiple responses

### Factors associated with HIV-self testing among medical/nursing students

After controlling for confounders, the following factors were significantly associated with self-testing for HIV: age, program of study, year of study, religion, number of sexual partners, and inconsistent use of condoms (table 4). Increase in age significantly reduced the odds of HIV self-testing by 11%. Students who were pursuing the BNA program were significantly less likely to test for HIV at home compared to students who were pursuing the MBChB program. Consistent use of condoms and Muslim religion significantly reduced the odds of self-testing for HIV. Students in year four of study compared to those in year five, and having 2 or more partners in the last year increased the odds of HIV self-testing.

**Table 4:** Bivariate and multivariate logistic regression for factors associated with HIV self-testing.

| Variable | COR (95% CI) | P-value | AOR (95% CI) | P-Value |
| --- | --- | --- | --- | --- |
| **Age (continuous variable)** | 0.97 (0.92-1.01) | 0.123 | 0.89(0.80-0.98) | 0.018 |
| **Gender** |  | 0.073 |  |  |
| Male | Ref |  | Ref |  |
| Female | 0.63 (0.38-1.04) |  | 0.66(0.32-1.34) | 0.249 |
| **Program of study** |  |  |  |  |
| MBChB | Ref |  | Ref |  |
| BNS | 1.37 (0.73-2.55) | 0.326 | 1.24 (0.51-3.02) | 0.638 |
| BNA | 0.89 (0.87-1.61) | 0.741 | 0.40 (0.17-0.95) | 0.039 |
| **Year of study** |  |  |  |  |
| 1 | 1.66 (0.71-3.88) | 0.240 | 0.61 (0.18-2.03) | 0.423 |
| 2 | 1.82 (0.83-3.98) | 0.132 | 1.03 (0.36-2.98) | 0.955 |
| 3 | 1.21 (0.52-2.77) | 0.658 | 1.02 (0.34-3.01) | 0.974 |
| 4 | 2.57 (1.17-5.65) | 0.019 | 2.90 (1.01-8.31) | 0.048 |
| 5 | Ref |  | Ref |  |
| **Marital status** |  |  |  |  |
| Married/cohabiting | Ref |  | Ref |  |
| Single | 1.16 (0.71-1.89) | 0.560 | 1.29 (0.60-2.77) | 0.517 |
| **Religion** |  |  |  |  |
| Christian | Ref |  | Ref |  |
| Muslim | 0.19 (0.04-0.91) | 0.038 | 0.07 (0.01-0.41) | 0.003 |
| **Employment** |  |  |  |  |
| Yes | Ref |  | Ref |  |
| No | 1.09 (0.65-1.83) | 0.734 | 0.45 (0.18-1.16) | 0.101 |
| **Age at sex debut (continuous variable)** | 0.94 (0.85-1.03) | 0.179 | 1.02 (0.89-1.19) | 0.720 |
| **HIV testing at sex debut** |  |  |  |  |
| Yes | Ref |  | Ref |  |
| No | 1.85 (1.12-3.06) | 0.016 | 1.49 (0.69-3.22) | 0.307 |
| **Number of sex partners in the past year** |  |  |  |  |
| $\geq 2$ | 2.32 (1.40-3.85) | | 3.22 (1.49-7.00) | |
| 1 | Ref | 0.001 | Ref | 0.003 |
| **Disclosed HIV status** |  |  |  |  |
| Yes | Ref |  | Ref |  |
| No | 0.59 (0.35-0.98) | 0.044 | 1.02 (0.47-2.22) | 0.961 |
| **Inconsistent use of condoms** |  |  |  |  |
| Yes | Ref |  | Ref |  |
| No | 0.65 (0.34-1.26) | 0.202 | 0.36 (0.15-0.88) | 0.024 |
| **Broken condoms** |  |  |  |  |
| Yes | Ref |  | Ref |  |
| No | 1.32 (0.81-2.17) | 0.267 | 1.59 (0.74-3.38) | 0.232 |
| **Sex with partner of unknown status** |  |  |  |  |
| Yes | Ref |  | Ref |  |
| No | 0.56 (0.33-0.97) | 0.040 | 1.29 (0.53-3.10) | 0.572 |
| **Frequency of HIV testing in the past year** |  |  |  |  |
| 0 | Ref |  | Ref |  |
| 1 | 2.66 (0.72- 9.83) | 0.143 | 3.92 (0.86-17.96) | 0.078 |
| $\geq 2$ | 2.94 (0.87-9.86) | 0.081 | 3.51 (0.87-14.10) | 0.077 |

## Discussion

The study was conducted to determine HIV testing preferences, uptake of HIVST and its associated factors among young adults in a public university. Overall, 55% of the young adults had tested by themselves at home which was congruent with their preferences to use HIV self-testing in the future. HIV self-testing was preferred over provider-based HIV testing because of the associated privacy and convenience linked to HIVST. These study findings should be interpreted in light of its limitations. Although we maintained anonymity in data collection, social desirability bias is likely given the sensitivity of the topic under study.

Nevertheless, because we measured for both the preferences and actual behaviours in HIV self-testing, the study findings are likely to be valid. The study provides important implications that can promote adoption of HIV testing approaches that can improve HIV testing among young adults. Ultimately, the improved HIV testing can contribute to the attainment of the sustainable development goals to reduce the AIDS epidemic by 2030 [3].

In our study, the uptake of HIV self-testing was 55% which was consistent with 56% of participants who preferred to use HIV self-testing over provider-based in the future among young adults. This highlights a congruence between preferences for HIV self-testing among young adults and their actual behaviours. The HIV self-testing in our study was higher than 24% reported among university students in Central Uganda [11]. The high uptake of HIV self-testing in our study may be attributable to the study population who were medical and nursing students, and thus, had some level of knowledge regarding HIV testing.

The majority of young adults preferred HIV self-testing which was consistent with high preferences and acceptability for HIVST in most of the studies [11,13,14]. Qualitative studies have equally reported that young adults find HIVST services to be more preferable, acceptable, feasible and appropriate [10,12]. Contrary to our study findings, 77% of adolescents and young girls in Kenya preferred provider-aided HIV testing to HIVST probably related to low levels of literacy [15]. This highlights that one size may not fit all, and that it is critical to consider the socio-demographic profile of young adults especially their levels of literacy in promoting HIVST practices.

One of the major findings in our study was that young adults valued privacy and confidentiality associated with HIV self-testing. Previous studies have equally reported that young adults prefer HIV self-testing because of its privacy and confidentiality [10,16]. HIVST eliminates youth-related barriers that affect uptake of HIV testing services among young adults [10,12]. Young adults face numerous barriers such as provider bias, discrimination, stigma, and negative judgement from older clients in the clinic-based HIV testing all of which were not of much concern in HIVST [10]. Adolescents and young adults are able to test themselves for HIV in the privacy of their homes without judgment, discrimination or mistreatment in the health facilities [10]. The resulting confidentiality of the test results enables young adults to maintain their dignity and autonomy while accessing HIV testing services [10,13].

Previous studies have underscored that young adult find HIVST to be more convenient, easy to use, efficient, and that it saves time and money [16-18]. This was consistent with our study findings where young adults preferred HIVST and the majority of them used HIVST because it was more convenient for them. The convenience in HIV self-testing can enable couples to test before sex and promote safer sex practices, while it is also known to increase HIV testing across all populations including young adults [9]. The convenient self-testing can assuage the lack of youth-friendly health services which deters the majority of young adults from going to health facilities for HIV testing [10]. Furthermore, convenience in self-testing for HIV eliminates health system bottlenecks especially in a setting like Uganda where long waiting times, distant health facilities, lack of HIV testing services, and shortage of personnel can deter many young adults from testing for HIV [17].

Young adults in our study who preferred provider-based HIV testing over HIVST did so because testing in the health facilities were seen to provide more accurate, trustworthy, credible, and reliable test results. Although the accuracy of test results in HIVST is equivalent to that from health facility-based HIV testing [15,19], previous studies have underscored young adults’ scepticism over the accuracy of test results obtained from HIVST [18,20]. Concerns over accuracy in HIVST were even more pronounced in oral-based HIV test kits than in blood-based HIV test kits [21]. Therefore, it is important to address fears and scepticism regarding accuracy of results from HIVST as concerns over test accuracy may act as a deterrent for use of HIVST among young adults [17].

Previous studies have highlighted the lack of pre-test and post-test counselling in HIV testing as a major concern for individuals seeking HIV testing [14,17]. Our study findings were consistent with studies which cited lack of counselling as a major deterrent for uptake of HIVST [10,20]. The lack of counselling in HIVST has been suggested to result in gender-based violence, emotional distress, suicide and self-harm though evidence to that effect is currently lacking [14,18,20]. This may be because individuals who opt for HIV self-testing are informed that HIVST does not confirm the HIV status but that it serves to screen for HIV [9]. HIVST approaches that include virtual counselling by healthcare providers through the mobile telephone have been suggested to meet the needs of young adults to access counselling before and after the HIV test [21]. Post-test counselling over the telephone can reduce maladaptive behaviours in case of the positive test [21].

The predictors for uptake of HIVST in our study were consistent with those in the literature [11,20,22]. Participants with multiple sex partners were more likely to have used HIVST, a finding which was similar to that in Tanzania [20]. The increased odds of HIVST among participants with multiple sex partners was plausible because the new partners may have to be tested at the point of sexual encounter. Our study was consistent with a study in Nigeria where young adults were less likely to have HIVST as they get older [22]. The increased likelihood of HIVST among younger people is likely because of higher sexual activity among younger people than their older counterparts. Similar to our study findings, previous studies have reported socio-demographic variables such as religion, program of study, and year of study to be associated with HIVST [11,20].

## Conclusion

Young adults preferred HIV self-testing over the provider-based HIV testing in the health facility. Privacy, confidentiality and convenience were the most common reasons given for preferring HIVST over clinic-based HIV testing. The preference for clinic-based HIV testing was because of counselling and the perception that the results in the health facilities were credible, accurate, and trustworthy. HIVST was associated with having multiple sex partners, Muslim religion, students doing anaesthesia program and those in the fourth year of study. Addressing fears of test accuracy in the HIVST, and the lack of counselling can promote increased preference to use HIVST which has wider implications on uptake of HIV testing, linkage to care, viral load suppression and reduced AIDS-related mortality.

## Data Availability

All relevant data are within the manuscript and its Supporting Information files.

## Abbreviations

HIV: Human Immunodeficiency syndrome
AIDS: Acquired immunodeficiency syndrome
HIVST: HIV self-testing
PLHIV: People living with HIV
HAART: Highly active antiretroviral therapy
CI: Confidence intervals
BNA: Bachelor of Anaesthesia
BSN: Bachelor of Science in Nursing
MBChB: Bachelor of Medicine and Surgery
MRRH: Mbale Regional Referral Hospital
REC: 

## Consent for publication

All authors have provided consent for publication

## Availability of data and materials

The datasets used and/or analyzed during the current study are available in the supplementary files.

## Author’s contributions

WN &JE participated in the conceptualization of the research idea, developed the study protocol, and obtained ethical clearance for the study. WN and GN participated in data collection and analysis. WN wrote the first draft of the manuscript while RCN, and JE reviewed the first draft of the manuscript. All the authors met the criteria for authorship and have approved the final draft for publication.

## Acknowledgment

We thank the study participants and the faculty in the Department of Nursing, Busitema University for their respective roles in the implementation of this study. We thank Ms. Madeline Powers for editing the manuscript.

